# Real-World Data of Tenecteplase Versus Alteplase in the Treatment of Acute Ischemic Stroke: A Single-Center Analysis

**DOI:** 10.1101/2024.01.02.24300742

**Authors:** Yu Yao, Xiaoqin Zhang, Chang Liu, Lingling Cai, Yisha Ying, Yuefei Wu, Jianhong Yang

## Abstract

**BACKGROUND AND PURPOSE:** Tenecteplase was demonstrated pharmacological superiority over alteplase, potentially translating into clinical benefits. Numerous studies have confirmed that the effectiveness and safety of tenecteplase in acute ischemic stroke (AIS) treatment may not be inferior to that of alteplase, and it has potential workflow advantages. This study aimed to evaluate whether tenecteplase’s use in routine clinical practice has time management advantages and corresponding clinical benefits.

**METHODS:** The study included AIS patients treated with alteplase at the first affiliated Hospital of Ningbo University from January 2022 to February 2023, and those treated with tenecteplase from March 2023 to November 2023. We compared baseline clinical characteristics, key reperfusion therapy time indices (onset-to-treatment time [OTT], door-to-needle time [DNT], and door-to-puncture time [DPT]), and clinical outcomes (24-hour post-thrombolysis National Institutes of Health Stroke Scale [NIHSS] improvement, and intracranial hemorrhage incidence) between the groups using univariate analysis. We also assessed hospital stay durations and used binary logistic regression to examine tenecteplase’s association with DNT and DPT target times, NIHSS improvement, and intracranial hemorrhage.

**RESULTS:** 120 patients treated with tenecteplase and 144 with alteplase were included in the study. Baseline characteristics showed no significant differences in demographic data (sex and age), vascular risk factors (body mass index, hypertension, diabetes), baseline NIHSS, mRS, and bridging thrombectomy (P > 0.05). However, the tenecteplase group had a higher prevalence of hyperlipidemia (21.7% vs. 12.5%, P=0.047) and a lower incidence of atrial fibrillation (21.7% vs. 34%, P=0.027). Key time indices for AIS reperfusion therapy, such as OTT (133 vs. 163.72, P=0.001), DNT (36.5 vs. 50, P < 0.001), and DPT (117 vs. 193, P=0.002), were significantly faster in the tenecteplase group. Additionally, the rates of DNT ≤ 45 min (65.83% vs. 40.44%, P < 0.001) and DPT ≤ 120 min (59.09% vs. 13.79%, P=0.001) were significantly higher in the tenecteplase group compared to the alteplase group. Tenecteplase was an independent predictor of achieving target times for DNT (OR 2.951, 95% CI 1.732-5.030; P < 0.001) and DPT (OR 7.867, 95% CI 1.290-47.991; P=0.025). Clinically, the proportion of patients with NIHSS improvement 24 hours post-thrombolysis was significantly higher in the tenecteplase group (64.17% vs. 50%, P=0.024). No significant differences were observed in the incidences of symptomatic intracranial hemorrhage (sICH) or any intracranial hemorrhage (ICH) (P > 0.05). Patients receiving tenecteplase had shorter hospital stays (6 vs. 8 days, P < 0.001). In binary logistic regression models, tenecteplase was an independent predictor of NIHSS improvement at 24 hours (OR 1.715, 95% CI 1.011-2.908; P=0.045). There was no significant association between thrombolytic choice and sICH or any ICH (P > 0.05).

**CONCLUSIONS:** Venous thrombolysis with tenecteplase in AIS treatment significantly reduced DNT and DPT. It was associated with early neurological function improvement (at 24 hours), without compromising safety compared to alteplase. Shorter length of hospital stays for patients were found in the tenecteplase group. The findings support tenecteplase’s application in AIS as a new treatment choice.

## BACKGROUND AND PURPOSE

Acute ischemic stroke (AIS) poses a significant public health challenge, adversely impacting the national economy and public welfare due to its high incidence, disability, mortality rates, and escalating medical expenses. Intravenous thrombolysis represents a safe and effective approach for AIS’s ultra-early treatment. Following the 1996 US FDA approval of the recombinant tissue plasminogen activator (alteplase, rt-PA), extensive researches have confirmed that alteplase can significantly improve clinical outcomes. However, its specificity for fibrin is moderate and the risk of intracranial hemorrhage still exists. Alteplase has a short half-life (4-5 min), and its administration is complex, requiring intravenous bolus followed by a continuous infusion for one hour; the efficiency of this ultra-early treatment workflow still needs improvement. Despite updates and iterations with alternatives such as recombinant human pro-urokinase (rhPro-UK), ancrod, and desmoteplase, a series of clinical trials have not demonstrated significant advantages in functional improvement. Moreover, these alternatives showed no significant difference, and in some cases, a slightly higher risk of hemorrhage, especially intracranial hemorrhage, compared to control drugs^[1-4]^. This was the case until the advent of tenecteplase (TNK-tPA). Tenecteplase, a DNA variant of alteplase ^[5]^, exhibits enhanced fibrin specificity and greater resistance to plasminogen activator inhibitor-1 (PAI-1) ^[6]^, effectively targeting thrombi. Its improved fibrin specificity minimizes systemic fibrinogen consumption, substantially reducing hemorrhage risk. Additionally, tenecteplase’s extended plasma half-life^[7, 8]^ permits a 5-10 second intravenous injection administration ^[9-11]^.

As a third-generation anti-fibrinolytic intravenous thrombolytic drug, tenecteplase boasts a well-characterized mechanism of action and significant practical advantages in administration, making it a promising candidate. Multiple studies have confirmed that its efficacy and safety may not be inferior to that of alteplase ^[12-17]^. And it has potential workflow advantages ^[18]^.

This study aims to evaluate whether tenecteplase’s use in routine clinical practice has time management advantages and corresponding clinical benefits, providing a basis for analyzing the rationale behind tenecteplase’s off-label application.

## METHODS

### Research subjects

This study included AIS patients who received alteplase at the first affiliated Hospital of Ningbo University from January 2022 to February 2023 and those treated with tenecteplase from March 2023 to November 2023. Eligible patients were: (1) 18 years or older; (2) diagnosed with ischemic stroke per established criteria, with measurable neurological deficits; (3) treated within 4.5 hours of symptom onset; (4) confirmed via CT or MRI to have no hemorrhage, extensive cerebral infarction, or other non-stroke pathologies; (5) provided informed consent, either personally or through family members^[19]^. Exclusion criteria included standard contraindications to alteplase. The administered dosages were 0.9mg/kg (maximum 90mg) for alteplase and 0.25mg/kg (maximum 25mg) for tenecteplase. In March 2023, ethical considerations for clinical tenecteplase use and off-label usage were thoroughly addressed, in compliance with relevant regulations (ethical review number 2023-03-59). This study received approval from the local medical ethics committee (ethics approval number 2023-175RS).

### Data collection

Clinical data were obtained from the emergency and inpatient information management system, including baseline characteristics (gender, age, body mass index [BMI], hypertension, diabetes, hyperlipidemia), baseline National Institutes of Health Stroke Scale (NIHSS; a 42-point scale that quantifies neurologic deficits in 11 categories, with higher scores indicating more severe deficits) scores, and baseline modified Rankin scale (mRS; a seven point ordered classification scale reflecting functional neurological outcomes from 0 to 6, with 0 indicating no symptoms of neurological deficits and 6 indicating death) scores. Additionally, post-thrombolysis 24-hour NIHSS scores, cerebral imaging results (CT or MRI was performed before treatment and 22 to 36 hours after thrombolysis treatment. Other CT scans were done if necessary.), and hospital stay durations were collected. Critical time points such as onset-to-treatment time (OTT), door-to-needle time (DNT), and door-to-puncture time (DPT) were calculated. Workflow outcomes focused on the percentage of patients treated within the recommended 45-minute DNT (as per international stroke guidelines) and the percentage receiving bridging thrombectomy within the 120-minute target DPT (set by the Stroke Prevention and Treatment Project Committee of the National Health Commission).

The workflow outcomes comprised the proportion of patients treated within the recommended 45-minute DNT as per international stroke guidelines ^[20, 21]^, and the proportion receiving bridging thrombectomy within the 120-minute DPT, the standard time frame established by the Stroke Prevention and Treatment Project Committee of the National Health Commission for advanced stroke centers.

Clinical outcomes encompassed both efficacy and safety measures. Efficacy was assessed by the improvement in the NIHSS score at 24 hours post-treatment. Safety outcomes included symptomatic intracranial hemorrhage (sICH), defined as any apparently extravascular blood in the brain or within the cranium that was associated with clinical deterioration, as defined by an increase of 4 points or more in the score on the NIHSS, or that led to death and that was identified as the predominant cause of the neurologic deterioration^[22]^, and any intracranial hemorrhage (ICH).

### Statistical Analysis

All statistical analyses were conducted using SPSS (version 23.0). Continuous data were presented as mean ± standard deviation or median (interquartile range), and categorical data as number (percentage). Univariate analysis was utilized to assess differences between groups. The Independent Student’s t-test was applied to normally distributed variables, and the Mann-Whitney test to non-normally distributed variables. Categorical variables were compared using Pearson’s chi-squared test or Fisher’s exact test. Variables that could potentially influence outcomes (age, hypertension, diabetes, baseline NIHSS, baseline mRS), those with a P-value < 0.05 in univariate analysis, and the type of thrombolytic drug were included in binary logistic regression models to identify independent predictors of outcomes. P-values < 0.05 were considered statistically significant.

## RESULTS

The study initially reviewed records of 124 patients treated with tenecteplase and 148 with alteplase. After 1 patient with incomplete data and 7 lost to follow-up being excluded, 120 patients treated with tenecteplase and 144 with alteplase were included in the study. The demographic data (male [75.8% vs. 68.1%, P=0.163] and age [66.5 vs. 67.75, P=0.122]) and vascular risk factors (BMI [23.75 vs. 23.92, P=0.727], hypertension [63.3% vs. 54.2%, P=0.133], diabetes [14.2% vs. 19.4%, P=0.256]) were comparable between groups. Significant differences were noted in the prevalence of hyperlipidemia (21.7% vs. 12.5%, P=0.047) and atrial fibrillation (21.7% vs. 34%, P=0.027) in the tenecteplase group. Baseline NIHSS (4.5 vs. 7, P=0.155) and mRS (3 vs. 3.5, P=0.633) scores, as well as the proportion undergoing bridging thrombectomy (20.0% vs. 22.9%, P=0.566), did not significantly differ (**Table 1**).

**Table 1.**
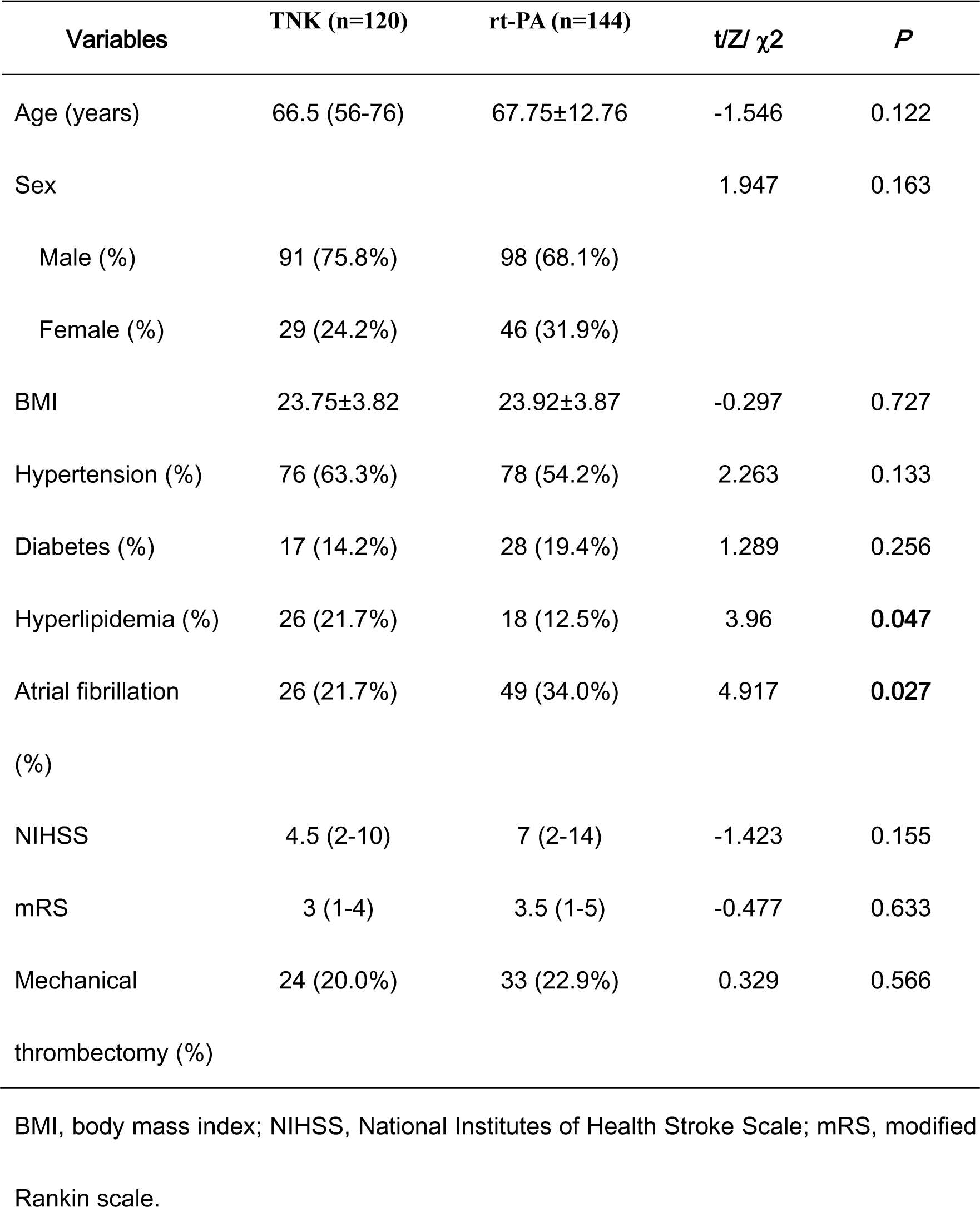
Characteristics of the Patients at Baseline.

In AIS reperfusion therapy, key time metrics such as OTT, DNT, and DPT were significantly lower in the tenecteplase group (OTT: 133 vs. 163.72, P=0.001; DNT: 36.5 vs. 50, P<0.001; DPT: 117 vs. 193, P=0.002). The proportions of patients in the tenecteplase group achieving DNT ≤ 45 min (65.83% vs 40.44%, P<0.001) and DPT ≤ 120 min (59.09% vs 13.79%, P=0.001) were significantly higher than in the alteplase group (**Table 2**). Binary logistic regression, incorporating baseline characteristics (age, hypertension, diabetes, hyperlipidemia, atrial fibrillation, baseline NIHSS, and baseline mRS) and thrombolytic drugs, indicated that tenecteplase was an independent predictor of meeting target times for DNT (OR 2.951, 95% CI 1.732-5.030; P<0.001) and DPT (OR 7.867, 95% CI 1.290-47.991; P=0.025) (**Table 4**). Baseline characteristics such as NIHSS did not affect workflow outcomes (P>0.05).

**Table 2.**
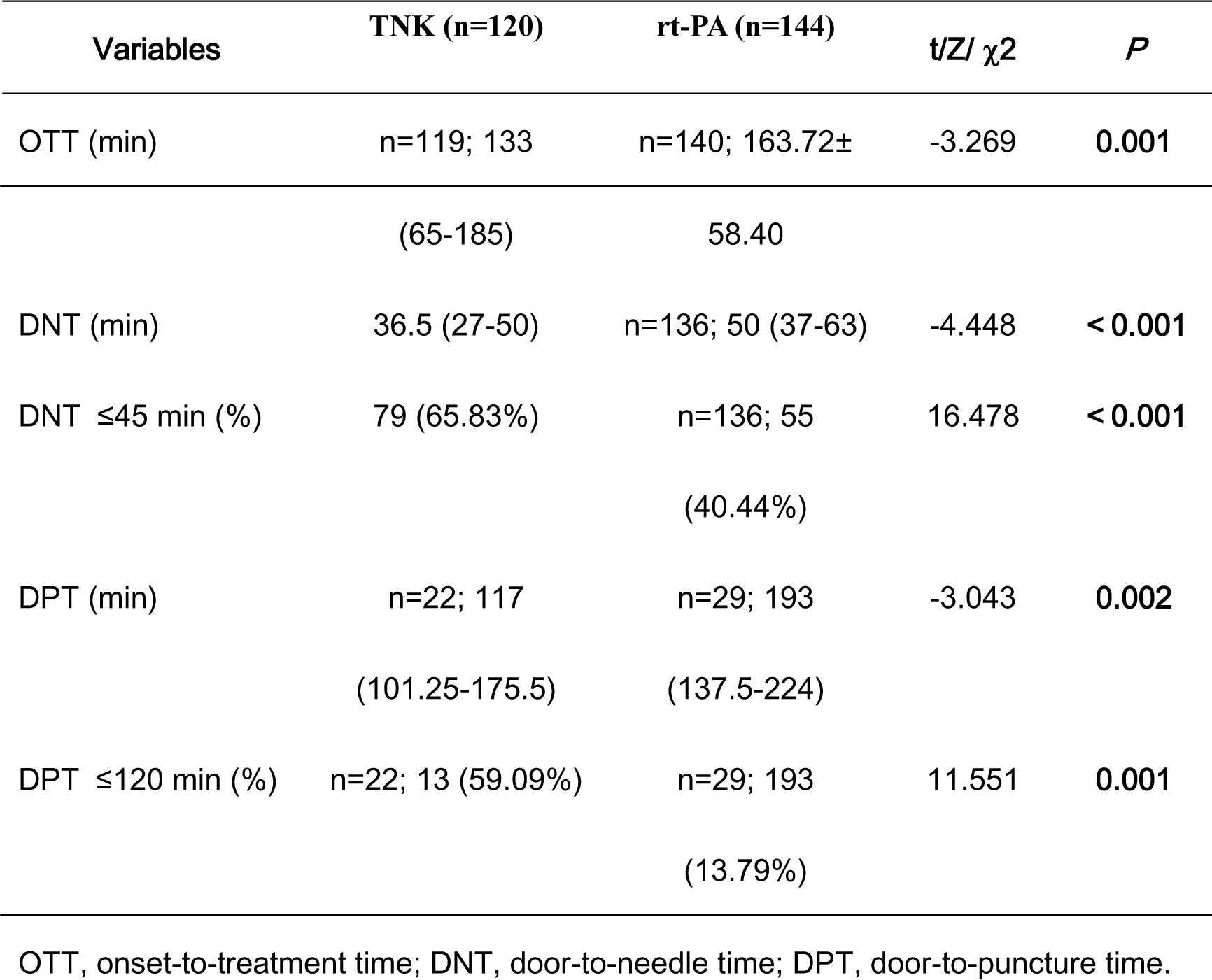
Treatment-Related Timings.

Regarding clinical outcomes, the tenecteplase group showed a significantly higher percentage of NIHSS improvement 24 hours post-thrombolysis (64.17% vs. 50%, P=0.024). The incidences of sICH (3.33% vs. 4.86%, P=0.536) or any ICH (13.3% vs. 15.28%, P=0.654) did not differ significantly. Tenecteplase patients had shorter hospital stays (6 vs. 8, P<0.001) (**Table 3**). Binary logistic regression revealed tenecteplase as an independent predictor of 24-hour NIHSS improvement (OR 1.715, 95% CI 1.011-2.908; P=0.045). Baseline NIHSS was identified as an independent risk factor for sICH (OR 1.082, 95% CI 1.020-1.147; P=0.009) and any ICH (OR 1.065, 95% CI 1.026-1.106; P=0.001). Atrial fibrillation was an independent risk factor for any ICH (OR 2.605, 95% CI 1.138-5.960; P=0.023). Other baseline characteristics and thrombolytic drugs did not significantly impact safety outcomes (**Table 5**).

**Table 3.**
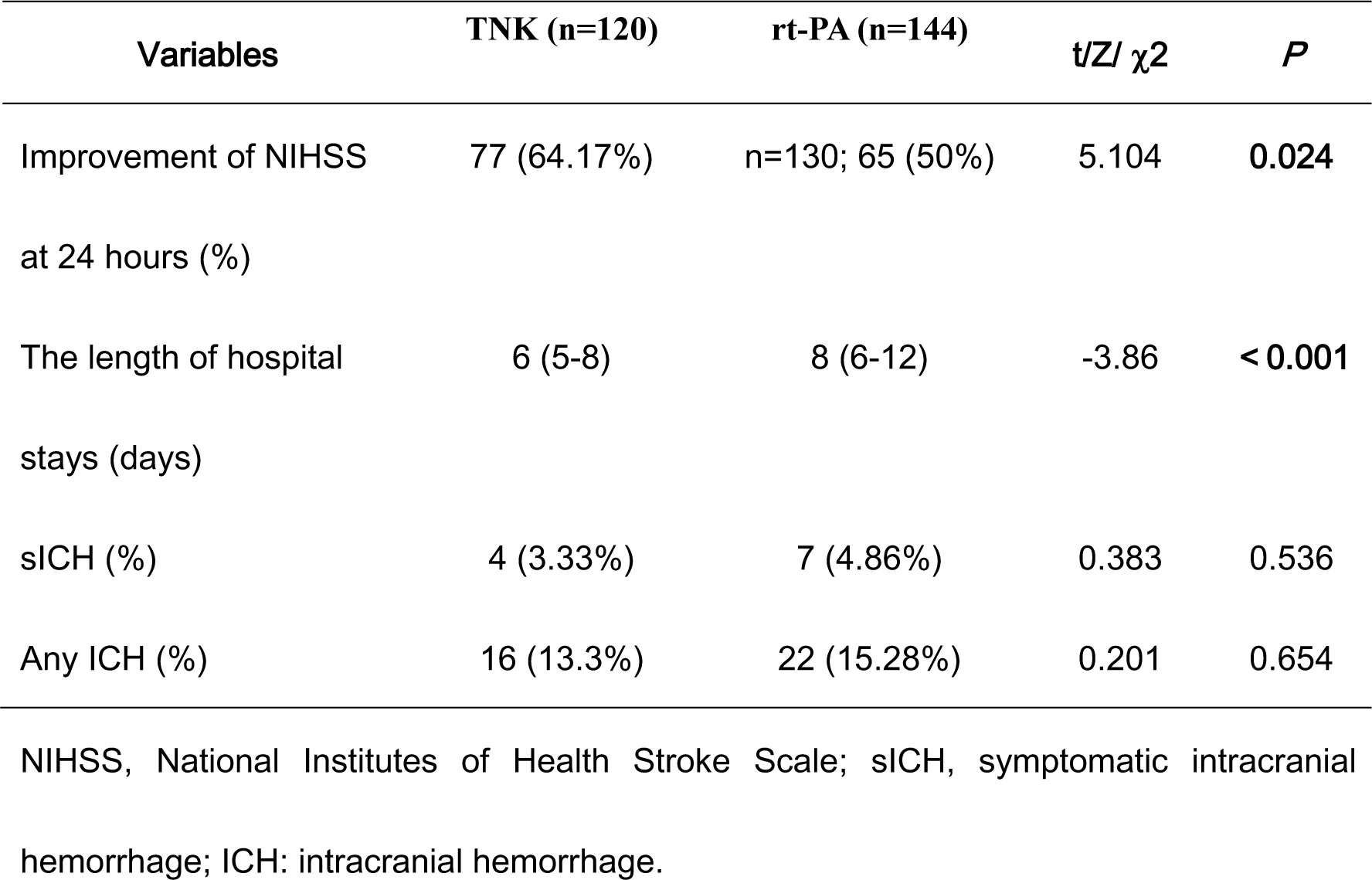
Univariate Analysis of Clinical Outcomes.

**Table 4.**
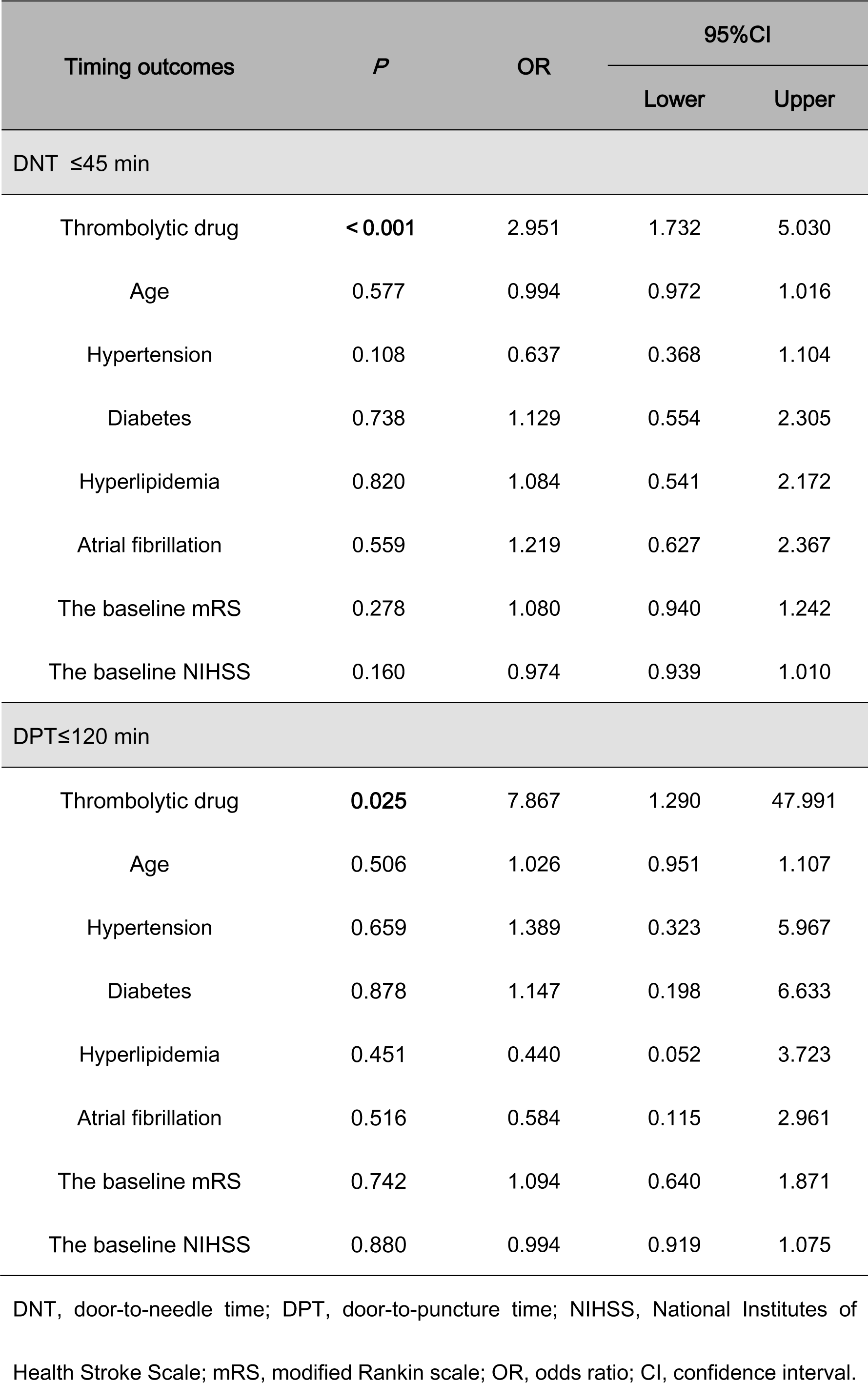
Multivariate Analysis of Workflow Outcomes.

**Table 5.**
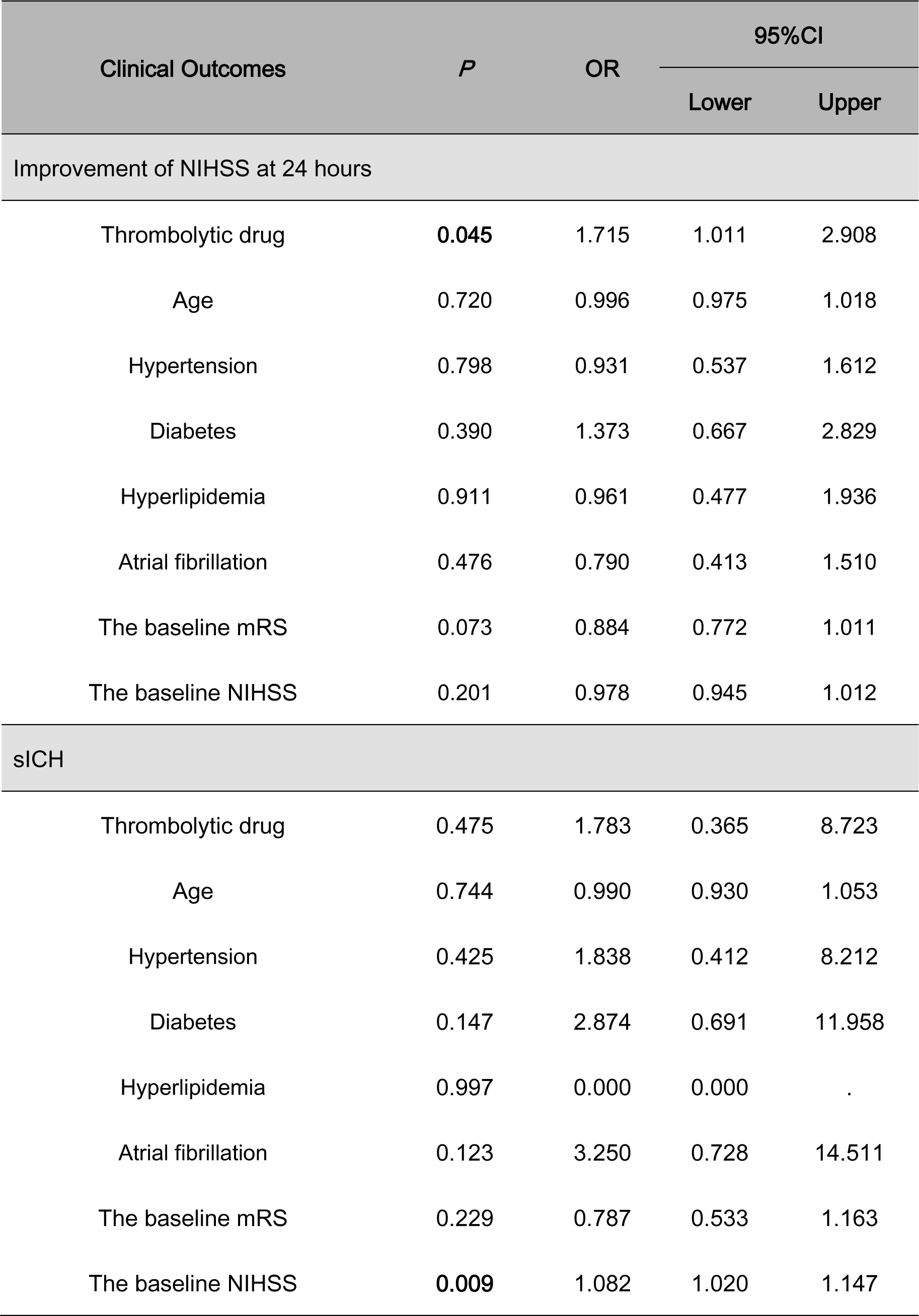
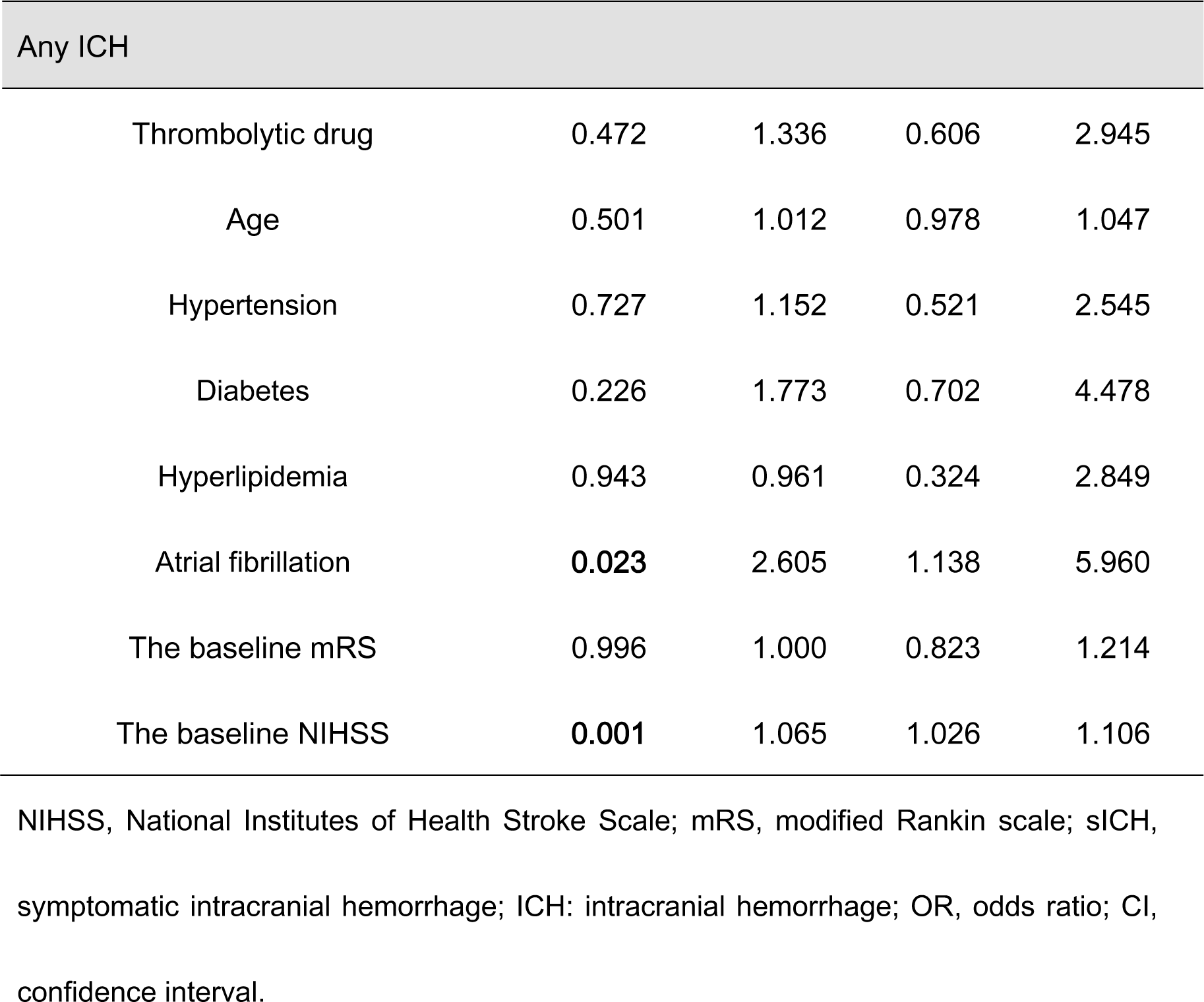
Multivariate Analysis of Clinical Outcomes.

## DISCUSSION

Our study is the first retrospective study on the application of tenecteplase in AIS in the real world from China. The study demonstrated that intravenous thrombolysis with tenecteplase is a feasible treatment for AIS, associated with early clinical improvement, enhanced ultra-early treatment workflow efficiency, and reduced hospital stays. Its safety outcomes paralleled those of alteplase.

Our study highlighted early neurological improvement in patients undergoing intravenous thrombolysis, identifying tenecteplase as an independent predictor of 24-hour NIHSS improvement, aligning with findings from the TASTE-A trial. Patients treated with tenecteplase at Melbourne mobile stroke units exhibited significantly smaller perfusion lesion volumes upon hospital arrival than those treated with alteplase, indicating a higher early reperfusion rate ^[13]^. Multiple studies in clinical practice have suggested that tenecteplase offers substantial clinical benefits. In AIS patients with acute large vessel occlusion (LVO), tenecteplase treatment resulted in higher initial angiography reperfusion rates ^[17]^, enhanced 24-hour reperfusion and clinical improvement ^[16]^, and improved 90-day functional outcomes ^[17]^. For all AIS patients eligible for thrombolysis, the tenecteplase group achieved non-inferior rates of long-term functional prognosis (mRS score of 0–1 at 90 days) ^[12, 14, 15]^. These benefits are attributable to tenecteplase’s optimized pharmacological properties. As a DNA variant of alteplase, tenecteplase undergoes molecular changes at three sites (T at site 103, N at site 117, and K at sites 296 to 299) ^[5]^, enhancing its specificity by 10-14 times compared to alteplase. It directly activates plasminogen into plasmin upon contact with thrombi and exhibits 80 times increased resistance to plasminogen activator inhibitor-1 (PAI-1) ^[6]^, thus effectively acting on thrombi with lower dosage but improved efficacy.

Regarding safety outcomes, the incidence of sICH (3.33% in the tenecteplase group and 4.86% in the alteplase group) and any ICH (13.33% in the tenecteplase group and 15.28% in the alteplase group) in the study was consistent with previous studies ^[14, 15, 22, 23]^. The incidences of sICH and any ICH were not significantly different between two groups, corroborating previous research ^[12-15]^. This safety profile is due to tenecteplase’s fibrin specificity ^[6]^, which minimizes systemic fibrinogen consumption and significantly reduces hemorrhage risk.

Supported by high-quality clinical trials, the use of tenecteplase in AIS treatment has been incorporated into several national AIS management guidelines ^[19, 20, 24]^. The 2022 European Stroke Organization Conference (ESOC) highlighted two register studies. Canadian researcher Katsanos, drawing on the international SITS-ISTR registry and the French multicenter TETRIS registry, demonstrated superior 90-day mRS score distributions, lower all-cause mortality, and no increased risk of sICH in the tenecteplase group. This supports the judicious use of tenecteplase in AIS treatment.

In addition to efficacy and safety, tenecteplase was found to enhance key time metrics in AIS treatment, aligning with recent prospective ^[18]^ and retrospective studies ^[30]^. Its half-life of 20-24 minutes enables a prolonged effective blood concentration ^[7, 8]^, allowing for a rapid 5-10 seconds intravenous injection, bypassing the need for infusion pumps required for alteplase’s hour-long infusion ^[9-11]^. Tenecteplase can quickly initiate treatment without the need for infusion pumps or additional equipment, significantly reducing DNT. The efficacy of intravenous thrombolysis is time-sensitive, with delays diminishing its benefits. DNT, a controllable hospital metric, is crucial for predicting the prognosis of AIS patients receiving thrombolysis ^[25, 26]^. It is also a key indicator for establishing efficient stroke pathways. For patients with large vessel occlusion (LVO), current guidelines recommend a bridging treatment approach combining intravenous thrombolysis and arterial embolectomy when criteria for both are met, rather than proceeding directly to endovascular treatment (EVT). Different from alteplase, tenecteplase does not require standard infusion pumps for a one-hour intravenous infusion. This facilitates quicker patient transfer after start of thrombolysis, enhancing stroke green channel management processes and significantly reducing DPT. EVT’s effectiveness is similarly time-dependent^[27]^. Shortening DPT and achieving prompt reperfusion of occluded vessels are linked to improved clinical outcomes ^[28]^.

Novelly, our study observed a shorter length of hospital stay for patients in the tenecteplase group compared to the alteplase group, suggesting potential savings in medical resources. This may be related to the higher proportion of early neurological function improvement in the tenecteplase group, which accelerates patients’ recovery. Tenecteplase, being less expensive, showed a greater net benefit in overall hospital cost analyses, primarily due to lower hospitalization costs ^[18]^. This may also be due to the shortened length of hospitalization. This reduction in drug costs is crucial in cost-benefit analysis and could potentially enhance the savings and quality-adjusted life years in the healthcare system.

However, our study has limitations. (1) It was based on data from a single center, and it is uncertain if similar results would be replicated in other centers or regions. (2) Thrombolytic treatment assignment was neither randomized nor blinded and, therefore, subject to biases in management decisions and outcome assessments. Fortunately, there was not much difference in the baseline characteristics, and we attempted to attenuate this limitation by binary logistic regression. (3) The clinical outcomes reported were early-stage, and further exploration is needed to assess long-term neurological function improvement. Yet, these early indicators have been shown to predict 90-day mRS scores ^[29]^, validating their use. (4) Most participants had mild to moderate stroke, the impact of tenecteplase on more severe stroke outcomes remains to be investigated. Future research will aim to expand the sample size and extend follow-up duration to provide more detailed data, such as 90-day mRS scores.

In conclusion, tenecteplase, as a new-generation thrombolytic drug, demonstrated a higher rate of early clinical improvement in AIS treatment and safety comparable to alteplase. Its ease of administration and management significantly enhanced target DNT and DPT achievement rates. Although confirmation in larger multicenter studies and ongoing randomized trials is needed, this study supports the use of tenecteplase in AIS intravenous thrombolysis.

## Data Availability

The data are not publicly available due to their containing information that could compromise the privacy of research participants.

## Acknowledgements

None.

